# Gaps in Essential Maternal Health Services during Pregnancy in Somaliland: Evidence from a National Composite Index Analysis

**DOI:** 10.64898/2026.05.06.26352578

**Authors:** Abdiasis Aden Omer, Hamda Jama Yousuf, Ahmed Abdirahman Farah, Bashiir Mohamed Mohamoud, Mohamed Hussein Egeh, Ali Ahmed Hussein

## Abstract

Ensuring the coverage of essential maternal health services during pregnancy is critical for reducing maternal morbidity and mortality. However, in low-resource settings such as Somaliland, the completeness of antenatal care remains a major challenge. This study aimed to assess the prevalence and determinants of suboptimal essential maternal health services among women in Somaliland. A cross-sectional study was conducted using data from the 2020 Somaliland Demographic and Health Survey (SLDHS). A total of 2,835 women were included in the analysis. A composite index was constructed based on key antenatal care components, including blood pressure measurement, urine testing, blood testing, iron supplementation, malaria prophylaxis, and deworming treatment. The outcome variable was categorized as suboptimal (1) and adequate (0). Multilevel logistic regression analysis was performed to identify factors associated with suboptimal maternal health services, accounting for the hierarchical structure of the data. The prevalence of suboptimal essential maternal health services was 59.9%, while only 40.1% of women received adequate services. Preventive interventions such as iron supplementation (28.5%), malaria prophylaxis (0.5%), and deworming (0.9%) were particularly low compared to routine screening services. Higher educational attainment and wealth status were significantly associated with lower odds of suboptimal care, while multiparity and regional disparities were associated with higher odds. Adequate antenatal care utilization was the strongest protective factor (AOR = 0.006; 95% CI: 0.002–0.018). Suboptimal maternal health service delivery remains a significant challenge in Somaliland. Improving maternal health outcomes requires not only increasing antenatal care coverage but also ensuring the completeness and quality of essential service components. Targeted interventions addressing socioeconomic and regional inequalities are crucial.

## Background

One important measure of the effectiveness of the health system and the well-being of mothers is the availability of necessary maternal health services during pregnancy. In order to improve health outcomes, encourage positive pregnancy experiences, and ensure early detection and management of pregnancy-related complications, the WHO recommends at least eight antenatal care contacts. It has been demonstrated that adequate use of these services considerably lowers morbidity and mortality among mothers and newborns (WHO, 2016). However, there are still differences in access and use, particularly in environments that are unstable and affected by conflict(1). Since its creation in the World Health Assembly in 2005, Universal Health Coverage (UHC) has grown in importance as a major objective of the global health system. Target 3.8 of the World Health Organization’s Sustainable Development Goals (SDG) Agenda includes it. UHC, which encompasses service, financial, and population coverage, ensures that everyone has access to the high-quality healthcare services they require without experiencing financial hardship. Since the majority of maternal deaths are avoidable with prompt, high-quality care, universal coverage of high-quality intrapartum care services, such as Emergency Obstetric Care (EmOC) and delivery with a skilled birth attendant (SBA), is essential to lowering maternal mortality (2-4).

Globally, rates of maternal and infant mortality remain high, especially in environments with limited resources. In 2017, for example, 810 women per day died from avoidable pregnancy and childbirth-related causes, and 94% of all maternal deaths were reported in low-resource settings, mostly in Southern Asia and Sub-Saharan Africa. Preventable and treatable causes, such as severe bleeding, infections, pre-eclampsia, eclampsia, unsafe abortion, delivery complications, and others, account for about 75% of all maternal deaths (5-7). A woman worldwide passes away from pregnancy-related complications every two minutes, but the majority of these deaths could be avoided with tried-and-true interventions. The international community is urging low- and middle-income nations to reaffirm their commitment to lowering maternal and child mortality rates by expanding access to maternal and neonatal health services in light of the impending deadline for achieving the Millennium Development Goals. Of the 7.7 million deaths in 2010 attributed to children under five, 3.1 million occurred during the first 28 days of life(8).

Many sub-Saharan African nations continue to have suboptimal facility delivery, which is a reflection of ongoing limitations in health systems and access. Similar low levels of facility-based delivery utilisation are reported in studies from Tanzania (80%), Ghana (75.6%), Nigeria (41%), Eritrea (24.6%), Chad (23%), and Ethiopia (24%). Long travel times to medical facilities, difficulties with transport, direct and indirect costs, a lack of qualified healthcare professionals, and worries about the standard and decency of care are all common obstacles. The maternal continuum of care is also disrupted by these obstacles. To support the well-being of mothers and newborns, the World Health Organization recommends at least four antenatal care (ANC) visits and structured postnatal care contacts; however, women who give birth at home are less likely to receive these crucial services (9, 10). Approximately 62% of maternal deaths globally occur in Sub-Saharan Africa, which continues to bear a disproportionate burden of maternal mortality. With 621 maternal deaths per 100,000 live births as of 2020, Somalia has one of the highest maternal mortality ratios in the world, making it one of the most difficult countries in this region. This concerning statistic is situated in the complicated environment of ongoing hostilities, frequent humanitarian crises, and weak health infrastructure, all of which make it more difficult to access and provide healthcare. Although Somalia has reduced maternal mortality by about 44%, from 1044 deaths per 100,000 live births in 2006 to 692 in 2020, these gains are still not enough to meet global targets. A major obstacle to reaching Sustainable Development Goal 3.1, which calls for lowering the global maternal mortality ratio to less than 70 per 100,000 live births by 2030, is the ongoing disparity in maternal healthcare utilisation, particularly antenatal care (11, 12).

Significant obstacles still exist in low-resource environments like Somaliland despite international efforts to improve maternal health. With limited access to skilled birth attendance, emergency obstetric services, and high-quality antenatal care (ANC), especially for rural and nomadic populations, Somaliland’s maternal health service utilisation remains suboptimal. The Somaliland Health and Demographic Survey (SHDS 2020) revealed that while a significant percentage of women attend at least one ANC visit, the percentage completing the recommended number of ANC contacts remains low, according to the Somaliland Ministry of Health Development and the Central Statistics Department of Somaliland (13). Furthermore, health system constraints such as shortages of skilled healthcare providers, inadequate infrastructure, and limited availability of essential maternal health services continue to hinder progress toward achieving Universal Health Coverage (UHC) in Somaliland. Strengthening maternal health service delivery and improving equitable access remain critical priorities to reduce maternal mortality and improve pregnancy outcomes in the region.

## Methods and materials

### Study setting

This study focused on Somaliland; a self-declared republic located in the Horn of Africa that has not received formal international recognition. The country comprises six administrative regions, Awdal, MaroodiJeex, Sahil, Togdheer, Sanaag, and Sool, and spans approximately 176,120 square kilometers, with an estimated population of 5.7 million in 2021. A major challenge facing Somaliland is the limited availability and accessibility of healthcare services, particularly in remote rural areas(14).

### Data source, study design, and sampling procedure

The 2020 Somaliland Demographic and Health Survey provided the data we utilized. It seeks to provide current data to track the nation’s health status. The SLDHS is a nationally representative survey to gather information on a wide range of health-related issues, such as access to healthcare, fertility rates, nutrition, HIV/AIDS, housing conditions, family planning, gender-based violence, female circumcision, and chronic illnesses. The SLHDS 2020 collected data from a variety of communities using a reliable, multi-stage stratified cluster sampling approach. The sample process was adapted to the geographic setting; nomadic regions used a two-stage design, while urban and rural areas used a three-stage stratified cluster approach. Using probability proportional to size (PPS) based on dwelling structures and household lists, respectively, primary sampling units (PSUs) and secondary sampling units (SSUs) were determined. To provide a solid and trustworthy dataset for analysis, households were methodically chosen as the ultimate sampling units(14, 15).

### Study variables

The dependent variable, suboptimal essential maternal health services during pregnancy, was constructed as a composite index based on key antenatal care components, including blood pressure measurement, urine testing, blood testing, iron supplementation, malaria prophylaxis, and deworming treatment. Each component was coded dichotomously as 1 if the service was not received and 0 if the service was received. These components were then combined to generate an overall index, which was further categorized into two groups: women who received all essential services were classified as having adequate maternal health services, whereas those who missed one or more components were classified as having suboptimal services. For regression analysis, the outcome variable was coded as 1 = suboptimal and 0 = adequate, allowing the model to estimate the likelihood of experiencing suboptimal maternal health services.

The independent variables were categorized into individual-level and community-level factors. Individual-level variables included maternal age, educational level, wealth index, parity, antenatal care utilization, employment status, and media exposure. Community-level variables comprised place of residence, region, and perceived distance to a health facility. The selection of these variables was informed by existing literature and the availability of relevant data within the SLDHS dataset.

### Data analysis

The data were analyzed using Stata version 17, accounting for the complex survey design of the Somaliland Demographic and Health Survey (SLDHS 2020). Sampling weights, clustering, and stratification were applied using the survey (svy) commands to obtain nationally representative estimates. Descriptive statistics were computed to summarize the socio-demographic and obstetric characteristics of the study population, and results were presented using frequencies and weighted percentages. Bivariate analysis was conducted to assess the association between each independent variable and the outcome variable using the design-adjusted Pearson chi-square test. Variables with a p-value less than 0.25 in the bivariate analysis were considered candidates for multivariable modeling. A multilevel logistic regression model was fitted to identify factors associated with suboptimal essential maternal health services, accounting for the hierarchical structure of the data, where individuals were nested within clusters. Four models were constructed: a null model without explanatory variables to assess the extent of clustering, a model including only individual-level variables, a model including only community-level variables, and a final model incorporating both individual- and community-level factors. The strength of association was reported using adjusted odds ratios (AORs) with 95% confidence intervals. Statistical significance was declared at a p-value less than 0.05. Model comparison was performed using the log-likelihood, Akaike Information Criterion (AIC), and Bayesian Information Criterion (BIC), with lower values indicating better model fit. The intraclass correlation coefficient (ICC) was used to quantify the proportion of total variance attributable to cluster-level differences. The model with the lowest AIC and BIC values was considered the best-fitting model.

## Result

A total of 2,835 women were included in the analysis. The majority of participants were aged 25–29 years (27.8%), followed by those aged 30–34 years (22.9%), while women aged 45– 49 years constituted the smallest proportion (2.1%). Most respondents had no formal education (78.1%), with only a small proportion attaining secondary (4.4%) and higher education (2.0%). In terms of socioeconomic status, 38.1% of women belonged to the lowest wealth quintile, whereas 18.7% were in the highest quintile. More than half of the participants (55.5%) resided in rural areas. Regionally, the highest proportions were observed in Sanaag (18.4%) and Sool (14.1%). A considerable proportion of women (61.6%) reported distance to a health facility as a major barrier to accessing care. Regarding antenatal care utilization, the vast majority (86.7%) had inadequate visits, while only 13.3% achieved adequate antenatal care. In terms of parity, most women were multiparous (43.2%) or grand multiparous (43.0%). Employment levels were extremely low, with only 0.6% of women reporting employment, and a substantial proportion (59.9%) had no exposure to mass media (Table 1).

**Table 1:** Weighted distribution of socio-demographic and obstetric characteristics of women in Somaliland (SLDHS 2020, n = 2,835).

| **Variable** | **Category** | **Frequency (n)** | **Weighted %** |
| --- | --- | --- | --- |
| **Age group** | 15–19 | 162 | 5.1 |
|  | 20–24 | 520 | 17.6 |
|  | 25–29 | 764 | 27.8 |
|  | 30–34 | 602 | 22.9 |
|  | 35–39 | 497 | 17.0 |
|  | 40–44 | 221 | 7.4 |
|  | 45–49 | 69 | 2.1 |
| **Education level** | No education | 2,362 | 78.1 |
|  | Primary | 369 | 15.5 |
|  | Secondary | 73 | 4.4 |
|  | Higher | 31 | 2.0 |
| **Wealth index** | Lowest | 1,079 | 38.06 |
|  | Second | 474 | 16.72 |
|  | Middle | 304 | 10.72 |
|  | Fourth | 447 | 15.77 |
|  | Highest | 531 | 18.73 |
| **Residence** | Rural | 1,415 | 55.5 |
|  | Urban | 1,420 | 44.5 |
| **Region** | Awdal | 365 | 9.4 |
|  | Maroodi-jeeh | 322 | 26.9 |
|  | Sahil | 334 | 5.3 |
|  | Togdheer | 476 | 25.9 |
|  | Sool | 653 | 14.1 |
|  | Sanaag | 685 | 18.4 |
| **Distance to health facility** | Big problem | 1,891 | 61.6 |
|  | Not a problem | 944 | 38.4 |
| **ANC visits** | Inadequate | 2,458 | 86.7 |
|  | Adequate | 377 | 13.3 |
| **Parity** | Primiparous | 362 | 13.8 |
|  | Multiparous | 1,217 | 43.2 |
|  | Grand multiparous | 1,256 | 43.0 |
| **Employment** | Yes | 11 | 0.6 |
|  | No | 2,824 | 99.4 |
| **Media exposure** | No | 1,829 | 59.9 |
|  | Yes | 1,006 | 40.1 |

The coverage of essential maternal health service components varied substantially. While blood pressure measurement was nearly universal (94.9%), other components showed considerable gaps. Only about six in ten women received urine (61.3%) and blood tests (61.2%).

In contrast, coverage of preventive interventions was extremely low, with only 28.5% of women receiving iron supplementation. Even more concerning, malaria prophylaxis (0.5%) and deworming treatment (0.9%) were almost absent. These findings indicate a substantial imbalance in the delivery of maternal health services, where basic screening is widely implemented but critical preventive interventions are largely neglected (Table 2).

**Table 2:** Coverage of essential maternal health service components during pregnancy in Somaliland (SLDHS 2020)

| Component | Received (%) | Not received (%) |
| --- | --- | --- |
| Blood pressure measurement | 94.9 | 5.1 |
| Urine test | 61.3 | 38.7 |
| Blood test | 61.2 | 38.8 |
| Iron supplementation | 28.5 | 70.3 |
| Malaria prophylaxis | 0.5 | 98.4 |
| Deworming treatment | 0.9 | 98.1 |

The prevalence of suboptimal essential maternal health services during pregnancy was high, with 59.9% of women experiencing suboptimal care, while only 40.1% received adequate services. As illustrated in Figure 1, this indicates that the majority of women did not receive all recommended components of antenatal care. This imbalance reflects significant gaps in the delivery of comprehensive maternal health services. The high burden of suboptimal care is consistent with deficiencies observed in specific service components, where basic screening services are more commonly delivered compared to essential preventive and treatment interventions. These findings suggest that improving maternal health outcomes in this setting requires not only increasing access to antenatal care but also ensuring the consistent provision of all essential service components.

**Figure 1:**
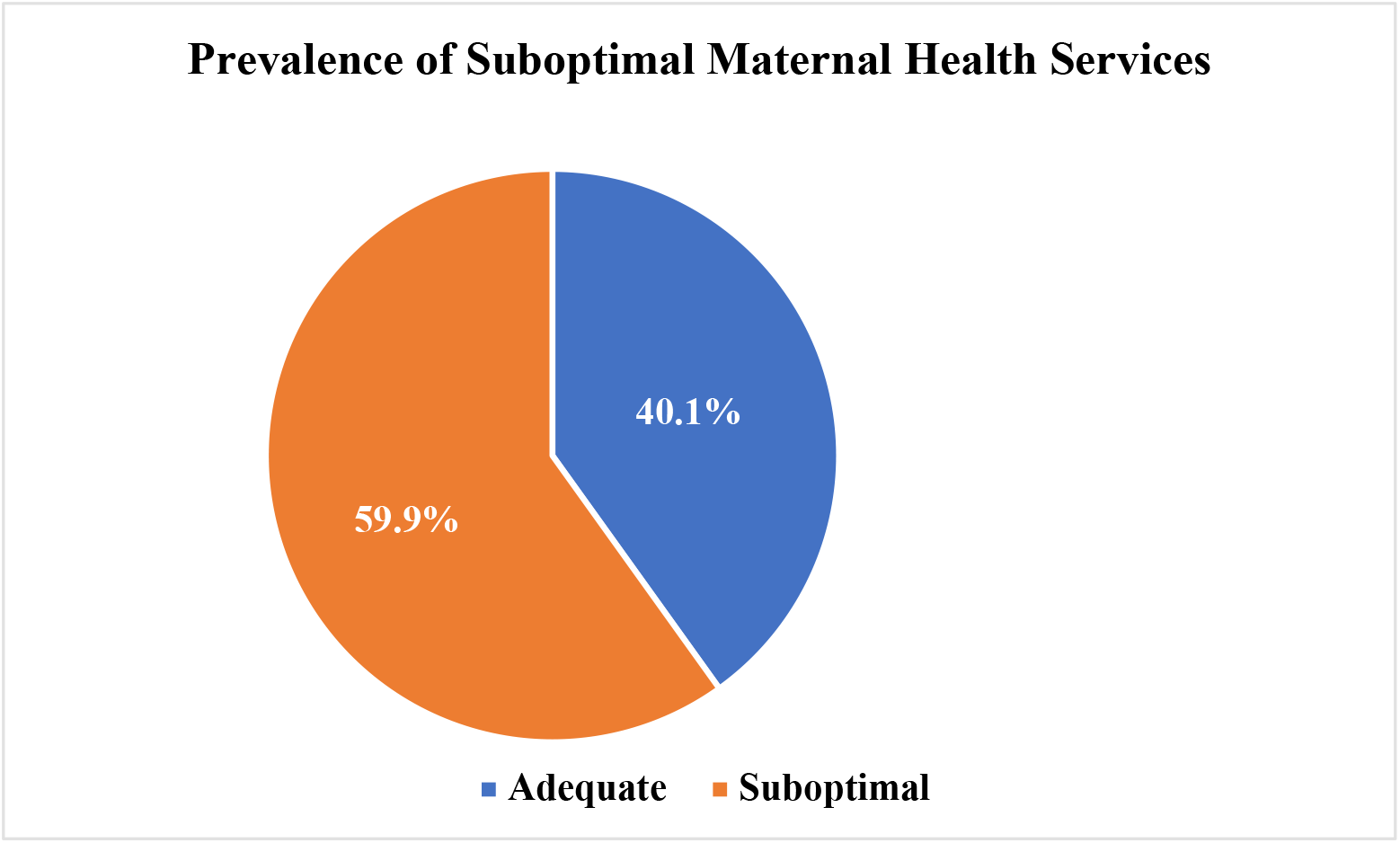
Distribution of adequate and suboptimal essential maternal health services among women in Somaliland (SLDHS 2020).

Bivariate analysis demonstrated that several socio-demographic and obstetric factors were significantly associated with suboptimal essential maternal health services. Maternal age showed a modest but statistically significant association (p = 0.034), with older women, particularly those aged 45–49 years, exhibiting higher proportions of suboptimal care (62.4%). Educational attainment displayed a strong inverse relationship with suboptimal care (p < 0.001), as women with no formal education had markedly higher levels of suboptimal care (56.5%) compared to those with higher education (1.9%). A clear socioeconomic gradient was also observed, with women in the lowest wealth quintile experiencing substantially higher suboptimal care (79.1%) compared to those in the highest quintile (17.7%) (p < 0.001). Significant regional disparities were evident (p < 0.001), particularly in Sool (72.6%) and Sanaag (71.9%), where the burden of suboptimal care was highest. Distance to a health facility was significantly associated with suboptimal care (p < 0.001), indicating the role of access-related barriers. Antenatal care utilization exhibited the strongest association (p < 0.001), with women who had inadequate visits showing a markedly higher prevalence of suboptimal care (59.5%), while nearly all women with adequate antenatal care received adequate services (99.7%). Parity was also significantly associated (p = 0.001), with grand multiparous women experiencing higher levels of suboptimal care (52.2%). In contrast, place of residence (p = 0.091) and employment status (p = 0.763) were not significantly associated with suboptimal maternal health services. Media exposure showed a significant association (p < 0.001), as women without media exposure had higher levels of suboptimal care (54.4%) compared to those with exposure (38.5%) (Table 3).

**Table 3:** Bivariate analysis of factors associated with suboptimal essential maternal health services among women in Somaliland (SLDHS 2020).

| Variable | Category | Adequate (%) | Suboptimal (%) | p-value |
| --- | --- | --- | --- | --- |
| Age group | 15–19 | 47.8 | 52.2 | **0.034** |
|  | 20–24 | 51.3 | 48.7 |  |
|  | 25–29 | 57.5 | 42.5 |  |
|  | 30–34 | 53.3 | 46.7 |  |
|  | 35–39 | 49.5 | 50.5 |  |
|  | 40–44 | 41.3 | 58.7 |  |
|  | 45–49 | 37.6 | 62.4 |  |
| Education | No education | 43.5 | 56.5 | **<0.001** |
|  | Primary | 76.8 | 23.2 |  |
|  | Secondary | 92.5 | 7.5 |  |
|  | Higher | 98.1 | 1.9 |  |
| Wealth index | Lowest | 20.9 | 79.1 | **<0.001** |
|  | Second | 34.0 | 66.0 |  |
|  | Middle | 52.1 | 47.9 |  |
|  | Fourth | 62.4 | 37.6 |  |
|  | Highest | 82.3 | 17.7 |  |
| Residence | Rural | 55.9 | 44.1 | 0.091 |
|  | Urban | 47.0 | 53.0 |  |
| **Region** | Awdal | 59.0 | 41.0 | **<0.001** |
|  | Maroodi-jeeh | 71.9 | 28.1 |  |
|  | Sahil | 50.8 | 49.2 |  |
|  | Togdheer | 59.4 | 40.6 |  |
|  | Sool | 27.4 | 72.6 |  |
|  | Sanaag | 28.1 | 71.9 |  |
| **Distance to health facility** | Big problem | 45.3 | 54.7 | **<0.001** |
|  | Not a problem | 62.7 | 37.3 |  |
| **ANC visits** | Inadequate | 40.6 | 59.5 | **<0.001** |
|  | Adequate | 99.7 | 0.3 |  |
| **Parity** | Primiparous | 64.2 | 35.8 | **0.001** |
|  | Multiparous | 52.3 | 47.7 |  |
|  | Grand multiparous | 47.8 | 52.2 |  |
| **Employment** | Yes | 59.4 | 40.6 | 0.763 |
|  | No | 51.9 | 48.1 |  |
| **Media exposure** | No | 45.6 | 54.4 | **<0.001** |
|  | Yes | 61.5 | 38.5 |  |

*Note: p-values are based on the Pearson chi-square test adjusted for survey design.*

Model comparison indicated substantial variation across clusters in the null model, as evidenced by a high intraclass correlation coefficient (ICC = 0.44), suggesting the appropriateness of multilevel modeling. The inclusion of individual-level variables in Model I markedly improved model fit, as reflected by a reduction in the log-likelihood value from −1671.47 to −1330.71 and a corresponding decrease in the ICC to 0.09, indicating that individual characteristics explained a considerable proportion of the variation. Model II, which incorporated community-level variables, also demonstrated improved model fit compared to the null model, although clustering remained relatively high (ICC = 0.41). The final model (Model III), which combined both individual- and community-level factors, exhibited the lowest AIC (2643.50) and BIC (2804.15) values, along with a substantially reduced ICC (0.04), indicating that it provided the best fit to the data. Furthermore, the likelihood ratio test confirmed that all models were statistically significant (p < 0.001), with Model III offering the most parsimonious and explanatory model (Table 4).

**Table 4:** Model fit statistics for multilevel models of suboptimal essential maternal health services in Somaliland (SLDHS 2020)

| Model | Log-likelihood | ICC | AIC | BIC | LR test | p-value |
| --- | --- | --- | --- | --- | --- | --- |
| **Null model** | -1671.47 | 0.44 | 3346.94 | 3358.84 | 475.45 | <0.001 |
| **Model I (Individual)** | -1330.71 | 0.09 | 2703.41 | 2828.36 | 36.04 | <0.001 |
| **Model II (Community)** | -1624.32 | 0.41 | 3264.64 | 3312.24 | 417.11 | <0.001 |
| **Model III (Full model)** | -1294.75 | 0.04 | 2643.50 | 2804.15 | 10.04 | <0.001 |

*Note: ICC = Intraclass Correlation Coefficient; AIC = Akaike Information Criterion; BIC = Bayesian Information Criterion*

Multilevel logistic regression analysis identified several significant determinants of suboptimal essential maternal health services. Maternal age was not significantly associated with suboptimal care across all models. In contrast, educational attainment demonstrated a strong and consistent protective effect. Women with primary (AOR = 0.38, 95% CI: 0.28–0.51), secondary (AOR = 0.23, 95% CI: 0.11– 0.46), and higher education (AOR = 0.11, 95% CI: 0.02–0.54) had significantly lower odds of experiencing suboptimal care compared to those with no formal education. Parity was also an important factor, as multiparous women had higher odds of suboptimal care (AOR = 1.65, 95% CI: 1.18–2.30), although the association for grand multiparous women was not statistically significant in the final model. A clear socioeconomic gradient was observed, with women in higher wealth quintiles demonstrating significantly reduced odds of suboptimal care, particularly those in the highest quintile (AOR = 0.17, 95% CI: 0.12–0.24). Antenatal care utilization emerged as the strongest determinant, with women who received adequate antenatal care showing a substantial reduction in the likelihood of suboptimal care (AOR = 0.006, 95% CI: 0.002–0.018). Distance to a health facility was also significantly associated with suboptimal care, although the direction of the association suggests a need for cautious interpretation. Furthermore, marked regional disparities were evident, with women residing in Sool (AOR = 2.74, 95% CI: 1.84–4.08) and Sanaag (AOR = 3.19, 95% CI: 2.13–4.78) experiencing significantly higher odds of suboptimal care compared to those in Awdal. Media exposure was not significantly associated with suboptimal care. Additionally, urban residence was associated with slightly increased odds of suboptimal care (AOR = 1.26, 95% CI: 1.02–1.56). Overall, both individual- and community-level factors played a significant role in shaping the adequacy of maternal health services (Table 5).

**Table 5:**
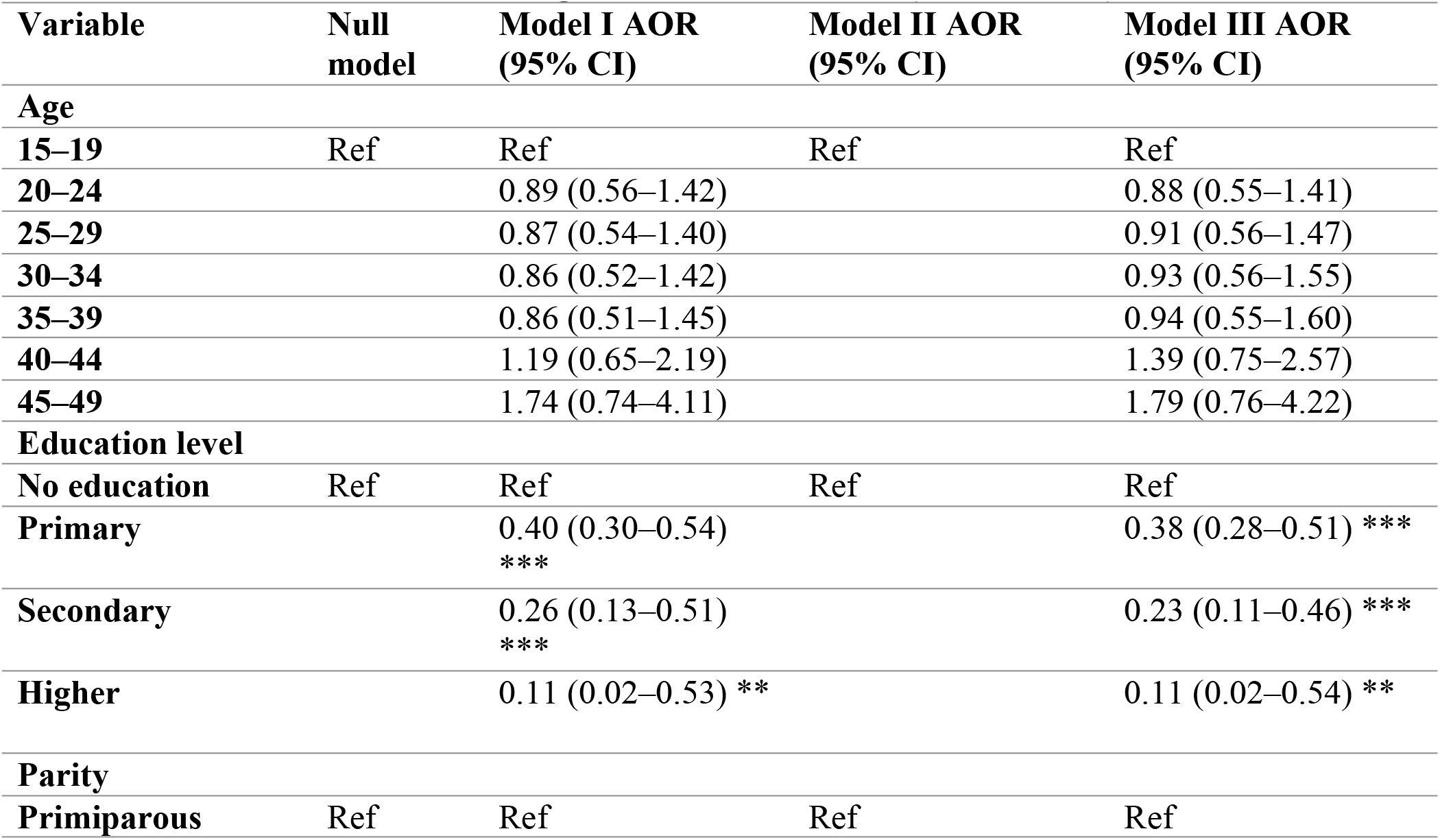
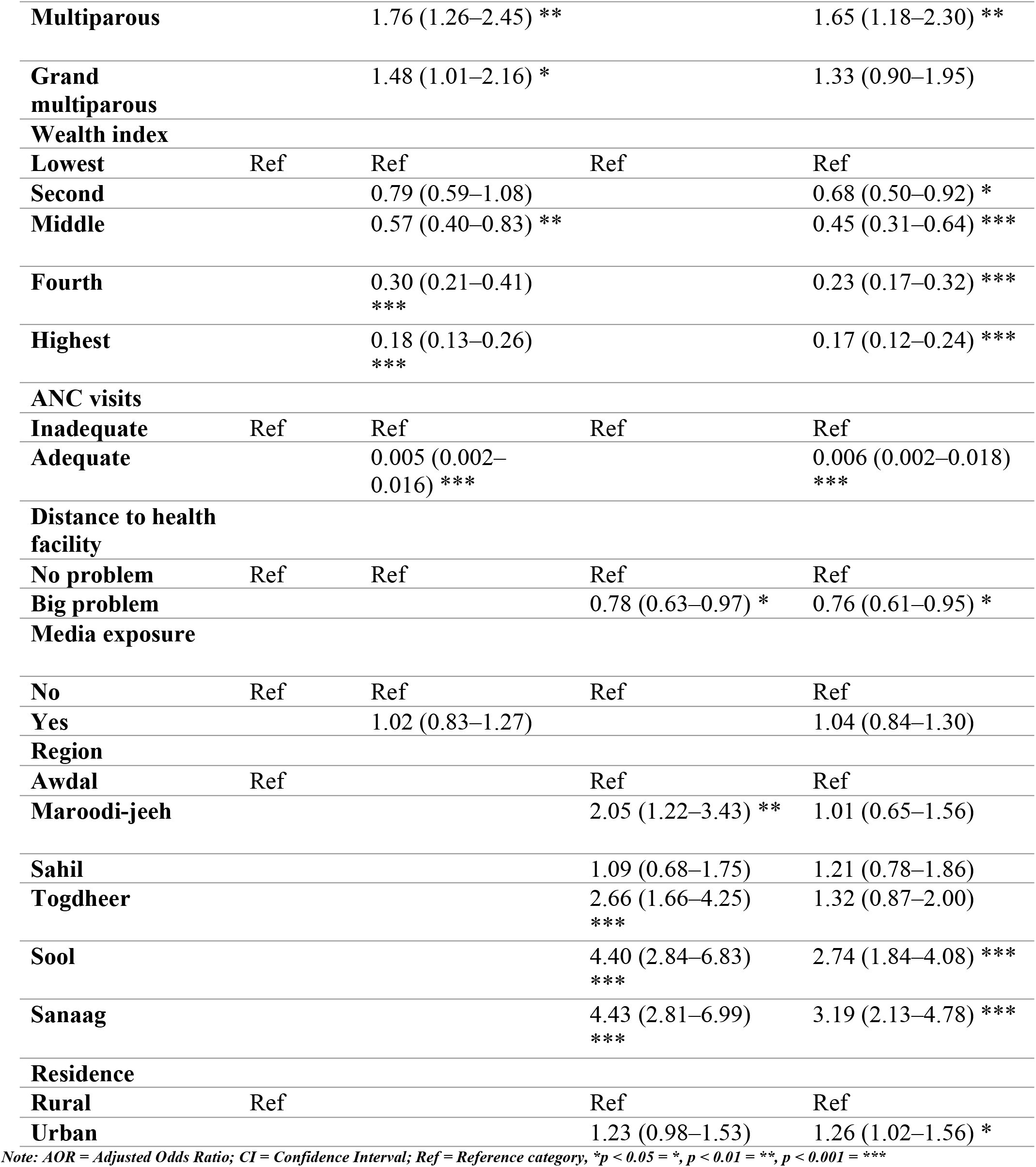
Multilevel logistic regression models of factors associated with suboptimal essential maternal health services among women in Somaliland (SLDHS 2020)

## Discussion

The findings of this study revealed that the prevalence of suboptimal essential maternal health services among women in Somaliland was high, with 59.9% experiencing suboptimal care, while only 40.1% received adequate services. This indicates substantial gaps in the delivery of comprehensive antenatal care services and highlights that access to care does not necessarily guarantee the receipt of all essential maternal health interventions. The prevalence of home birth among women who had utilized antenatal care services prior to delivery is 46.2%, with 14.7% and 31.5% in suboptimal and optimal ANC visits, respectively. The difference in proportion is statistically significant, and this implies that utilization of ANC in the two groups does not translate to utilization of health facility delivery, and hence highlights the major hitches in reducing maternal and child mortality in Nigeria(16-18). This agrees with findings from previous studies on ANC barriers in Nigeria, women’s attitude towards ANC service utilization in Ethiopia, and whether ANC translates to skilled birth attendants’ utilization in Ghana. The high prevalence of suboptimal care observed in this study can be partly explained by the uneven coverage of specific service components. While basic screening services such as blood pressure measurement were widely implemented, other essential interventions, including diagnostic tests and key preventive services such as iron supplementation, malaria prophylaxis, and deworming, were inadequately delivered. This imbalance suggests that the healthcare system prioritizes routine assessments over comprehensive service provision, resulting in incomplete maternal care. Similar patterns have been reported in other low- and middle-income countries, where gaps in service quality persist despite improvements in service contact.

Maternal education emerged as one of the strongest determinants of maternal health service adequacy. Women with higher levels of education were significantly less likely to experience suboptimal care, which is consistent with existing evidence indicating that education enhances health literacy, decision-making autonomy, and healthcare-seeking behavior. This aligns with previous results in low- and middle-income countries (LMIC)(19-22). Educated women are more likely to recognize the importance of antenatal care and to utilize available services effectively. This underscores the importance of investing in female education as a long-term strategy to improve maternal health outcomes. Socioeconomic status also played a significant role, with women from wealthier households demonstrating lower odds of suboptimal care. This finding reflects persistent inequalities in access to quality healthcare services (22-24). Financial constraints may limit the ability of poorer women to seek care, afford transportation, or access essential interventions, even when services are available. Addressing these disparities requires targeted policies aimed at reducing economic barriers and promoting equitable access to maternal healthcare. Antenatal care utilization was identified as the most influential factor, with women who received adequate antenatal care showing markedly reduced odds of suboptimal maternal health services. This finding highlights the critical role of ANC as a platform for delivering essential maternal health interventions (22). However, the low proportion of women achieving adequate ANC utilization indicates a dual challenge: both insufficient service uptake and incomplete service delivery among those who access care. Strengthening ANC coverage and ensuring adherence to recommended care protocols are therefore essential. Parity was also associated with suboptimal care, with multiparous women being more likely to experience inadequate services (25). This may be due to reduced perceived need for care among women with previous childbirth experience, leading to lower utilization of antenatal services. This finding emphasizes the need for targeted health education strategies that reinforce the importance of antenatal care for all pregnancies, regardless of prior experience.

Significant regional disparities were observed, particularly in Sool and Sanaag, where women had substantially higher odds of suboptimal care. These differences likely reflect variations in healthcare infrastructure, resource allocation, and geographic accessibility corroborated by previous studies in Nigeria (18, 24). Such disparities highlight the need for region-specific interventions aimed at strengthening health systems and improving service delivery in underserved areas. Interestingly, distance to a health facility was found to be significantly associated with suboptimal care, although the direction of the association was not entirely consistent with expectations(22). This may reflect differences in perceived versus actual access to healthcare, or the influence of unmeasured contextual factors. Therefore, this finding should be interpreted with caution and warrants further investigation. In contrast, maternal age, employment status, and media exposure were not significantly associated with suboptimal maternal health services in the final model. This suggests that structural and socioeconomic factors play a more prominent role than individual demographic characteristics in determining maternal healthcare outcomes in this context. Overall, the findings of this study highlight a “double burden” in maternal healthcare in Somaliland: low utilization of antenatal care services and inadequate delivery of essential care components. Addressing these challenges requires a comprehensive approach that strengthens both demand and supply sides of healthcare. Policy efforts should focus on improving female education, reducing socioeconomic inequalities, expanding access to antenatal care, and ensuring the consistent provision of all essential maternal health interventions across all regions.

## Methods and materials

### Study setting

This study focused on Somaliland; a self-declared republic located in the Horn of Africa that has not received formal international recognition. The country comprises six administrative regions, Awdal, MaroodiJeex, Sahil, Togdheer, Sanaag, and Sool, and spans approximately 176,120 square kilometers, with an estimated population of 5.7 million in 2021. A major challenge facing Somaliland is the limited availability and accessibility of healthcare services, particularly in remote rural areas(14).

### Data source, study design, and sampling procedure

The 2020 Somaliland Demographic and Health Survey provided the data we utilized. It seeks to provide current data to track the nation’s health status. The SLDHS is a nationally representative survey to gather information on a wide range of health-related issues, such as access to healthcare, fertility rates, nutrition, HIV/AIDS, housing conditions, family planning, gender-based violence, female circumcision, and chronic illnesses. The SLHDS 2020 collected data from a variety of communities using a reliable, multi-stage stratified cluster sampling approach. The sample process was adapted to the geographic setting; nomadic regions used a two-stage design, while urban and rural areas used a three-stage stratified cluster approach. Using probability proportional to size (PPS) based on dwelling structures and household lists, respectively, primary sampling units (PSUs) and secondary sampling units (SSUs) were determined. To provide a solid and trustworthy dataset for analysis, households were methodically chosen as the ultimate sampling units (USUs)(14, 15).

### Study variables

The dependent variable, suboptimal essential maternal health services during pregnancy, was constructed as a composite index based on key antenatal care components, including blood pressure measurement, urine testing, blood testing, iron supplementation, malaria prophylaxis, and deworming treatment. Each component was coded dichotomously as 1 if the service was not received and 0 if the service was received. These components were then combined to generate an overall index, which was further categorized into two groups: women who received all essential services were classified as having adequate maternal health services, whereas those who missed one or more components were classified as having suboptimal services. For regression analysis, the outcome variable was coded as 1 = suboptimal and 0 = adequate, allowing the model to estimate the likelihood of experiencing suboptimal maternal health services.

The independent variables were categorized into individual-level and community-level factors. Individual-level variables included maternal age, educational level, wealth index, parity, antenatal care utilization, employment status, and media exposure. Community-level variables comprised place of residence, region, and perceived distance to a health facility. The selection of these variables was informed by existing literature and the availability of relevant data within the SLDHS dataset.

### Data analysis

The data were analyzed using Stata version 17, accounting for the complex survey design of the Somaliland Demographic and Health Survey (SLDHS 2020). Sampling weights, clustering, and stratification were applied using the survey (svy) commands to obtain nationally representative estimates. Descriptive statistics were computed to summarize the socio-demographic and obstetric characteristics of the study population, and results were presented using frequencies and weighted percentages. Bivariate analysis was conducted to assess the association between each independent variable and the outcome variable using the design-adjusted Pearson chi-square test. Variables with a p-value less than 0.25 in the bivariate analysis were considered candidates for multivariable modeling. A multilevel logistic regression model was fitted to identify factors associated with suboptimal essential maternal health services, accounting for the hierarchical structure of the data, where individuals were nested within clusters. Four models were constructed: a null model without explanatory variables to assess the extent of clustering, a model including only individual-level variables, a model including only community-level variables, and a final model incorporating both individual- and community-level factors. The strength of association was reported using adjusted odds ratios (AORs) with 95% confidence intervals. Statistical significance was declared at a p-value less than 0.05. Model comparison was performed using the log-likelihood, Akaike Information Criterion (AIC), and Bayesian Information Criterion (BIC), with lower values indicating better model fit. The intraclass correlation coefficient (ICC) was used to quantify the proportion of total variance attributable to cluster-level differences. The model with the lowest AIC and BIC values was considered the best-fitting model.

## Conclusion

In conclusion, this study revealed a high prevalence of suboptimal essential maternal health services among women in Somaliland, indicating substantial gaps in the delivery of comprehensive antenatal care. Despite some progress in the provision of basic screening services, the coverage of critical preventive and treatment interventions remains inadequate. The findings demonstrate that access to antenatal care alone is insufficient to ensure optimal maternal health outcomes, as many women do not receive the full package of essential services. Key determinants such as maternal education, household wealth, antenatal care utilization, and regional disparities play a significant role in shaping the quality of maternal healthcare. These results highlight a critical need for health system strengthening that goes beyond increasing service coverage to improving the completeness and quality of care. Targeted interventions focusing on disadvantaged populations, particularly women with low education and those from poorer households, are essential. Furthermore, efforts should be made to enhance the effectiveness of antenatal care services by ensuring that all recommended components are consistently delivered across all regions. Addressing these gaps is crucial for improving maternal health outcomes and accelerating progress toward reducing maternal morbidity and mortality in Somaliland.

### Strengths and Limitations

This study has several important strengths. First, it utilized nationally representative data from the 2020 Somaliland Demographic and Health Survey, enhancing the generalizability of the findings to the wider population. Second, the use of a composite index to measure suboptimal essential maternal health services allowed for a more comprehensive assessment of service delivery, moving beyond single indicators to capture the completeness of antenatal care. Third, the application of multilevel logistic regression accounted for the hierarchical structure of the data, providing more robust estimates by incorporating both individual- and community-level factors.

However, this study is not without limitations. The cross-sectional design of the SLDHS limits the ability to establish causal relationships between the identified factors and suboptimal maternal health services. In addition, the reliance on self-reported data may introduce recall bias, particularly for events related to antenatal care services. Furthermore, some important variables, such as quality of care at health facilities and provider-level factors, were not available in the dataset and could not be included in the analysis. Finally, the construction of the composite outcome variable may inherently reflect components of antenatal care, which could partly explain the strong association observed between antenatal care utilization and the composite outcome variable.

### Policy Implications

The findings of this study have important implications for maternal health policy and program design in Somaliland. First, there is a need to shift focus from merely increasing antenatal care coverage to improving the quality and completeness of services delivered. Ensuring that all essential components of antenatal care, particularly preventive interventions such as iron supplementation, malaria prophylaxis, and deworming, are consistently provided should be a priority for health authorities. This requires strengthening clinical guidelines, improving provider training, and enhancing supervision and accountability mechanisms within health facilities.

Second, targeted interventions are needed to address socioeconomic and educational disparities in maternal healthcare. Women with low levels of education and those from poorer households were more likely to experience suboptimal care, highlighting the importance of pro-poor and equity-focused strategies. Policies such as conditional cash transfers, transportation support, and community-based outreach programs could help reduce financial and geographic barriers to accessing quality maternal health services.

Third, the significant regional disparities identified in this study call for region-specific strategies to improve service delivery in underserved areas, particularly in regions with higher burdens of suboptimal care. Strengthening health infrastructure, deploying skilled health workers, and expanding mobile or outreach services in hard-to-reach areas may help bridge these gaps.

Fourth, improving antenatal care utilization remains critical. Efforts should focus not only on increasing the number of visits but also on ensuring adherence to recommended care protocols during each visit. Integrating quality assurance mechanisms and routine monitoring of service components can help ensure that antenatal care visits translate into meaningful health outcomes.

Finally, strengthening health information systems is essential for monitoring the quality of maternal health services. Routine data collection and use should be enhanced to track the delivery of essential care components and identify gaps in real time, enabling timely and evidence-based decision-making.

## Data Availability Statement

The data used in this study are publicly available from online sources. Details of the data repository and access information can be found at: https://microdata.nbs.gov.so/index.php/catalog/50

## Ethics Statement

Ethical approval was not required for this study involving human participants, as it was conducted in accordance with applicable local regulations and institutional guidelines. The analysis was based on secondary data obtained from a publicly available dataset, and therefore, informed consent from participants or their legal guardians was not required, in line with national policies and institutional requirements.

## Author Contributions

AO: Conceptualization, Methodology, Formal analysis, and Writing – original draft. HY: Supervision, Methodology, and Writing – review and editing.

AF: Supervision, Visualization, and Writing – review and editing.

BM: Methodology, Formal analysis, Validation, and Writing – review and editing.

ME: Data curation, Formal analysis, Validation, and Writing – review and editing.

AH: Software and Writing – review and editing.

## Generative AI Statement

The authors declare that generative AI tools were used only for language editing and academic structuring support. All scientific content, analysis, and interpretations were developed and verified by the authors.

